# Role of Inflammation in the Pathogenesis of Anemia in Chronic Kidney Disease: Exploring the Impact of Inflammatory Biomarkers on Erythropoiesis and Erythropoietin Response

**DOI:** 10.1101/2025.01.01.25319873

**Authors:** Collince Odiwuor Ogolla, Lucy W. Karani, Stanslaus Musyoki

**Author notes:** **Corresponding Author:** Collince Odiwuor Ogolla.

## Abstract

**Background:** Anemia is one of the very common complications in Chronic Kidney Disease (CKD) and often included among patients who undergo hemodialysis. A very important aspect of anemia is that erythropoietin (EPO) resistance plays a very major role, whereas the role of inflammation to EPO resistance and impaired erythropoiesis is still poorly understood.

**Objective:** The objective of this study research therefore was to measure the effects of inflammation assessed by C-reactive protein (CRP) on anemia and in response to EPO in CKD patients undergoing hemodialysis here.

**Methods:** This is a cross-sectional study involving 120 CKD patients undergoing maintenance hemodialysis. Based on their CRP levels, participants were divided into two groups: High Inflammation (CRP ≥ 10 mg/L) and Low Inflammation (CRP < 10 mg/L). The hematological parameters such as hematocrit, serum ferritin, transferrin saturation, and EPO dose are evaluated. Statistical Analysis: Group comparisons and correlation analysis to study the relationships between CRP levels, iron status, and EPO resistance.

**Results:** In the high inflammation group, hematocrit levels (29.6 ± 4.8%) were significantly lower than in the low inflammation group in a comparison of (34.2 ± 5.2%, p < 0.001). The EPO dose was higher in the high inflammation group (3,200 ± 800 IU/week vs. 2,200 ± 700 IU/week, p < 0.001). Different parameters regarding iron, like serum ferritin and transferrin saturation, were significantly lesser in the high inflammation group. CRP levels had a negative correlation with hematocrit (r = -0.42, p < 0.001) and transferrin saturation (r = -0.36, p < 0.001), while they were found to have a positive correlation with the EPO dose administered (r = 0.48, p < 0.001).

**Conclusions:** Inflammation as measured by CRP has an important effect on anemia management in CKD patients undergoing hemodialysis.

## Introduction

Chronic Kidney Disease (CKD) is a common and severe complication with anemia, majorly seen in hemodialysis patients. It is mainly defined by the inhibited production of red blood cells due to decreased synthesis of erythropoietin (EPO), iron deficiency, and impairment in erythropoiesis defining.

Furthermore, inflammation plays an important role in pathophysiology regarding anemia with CKD, worsening the condition and increasing resistance towards EPO therapy. Gradual increases in inflammation as CKD progresses, such as an increase in CRP, have been associated with the severity of symptomatology, which further complicates anemia by interfering with iron metabolism and erythropoiesis (1, 2).

One of the main concerns in the management of anemia associated with CKD is erythropoietin resistance. Several patients fail to reach the target hematocrit levels despite receiving adequate dose of EPO, which in turn leads to a need for being high EPO doses, thereby increasing healthcare costs and adverse outcomes such as cardiovascular disease (3). It is believed that such resistance is mediated by several putative factors, one of which is inflammation that boosts pro-inflammatory cytokines production like interleukin-6 (IL-6) affecting negatively EPO responsiveness (4). Inflammation also elevates levels of hepcidin which is an important regulator of iron metabolism inhibiting iron absorption and utilization in erythropoiesis further complicating anemia management (5).

CRP used as a very well system failed marker in inflammation; however, it may be applied as a potential predictor of severity of anemia in a patient with CKD. Studies suggest that an increase in CRP would also relate to a decrease in hemoglobin and hematocrit values and could predict a poor response to EPO (6, 7). However, the exact mechanisms through which inflammation induces anemia and EPO resistance are not known as explicitly in CKD patients on hemodialysis. The present study was done to elicit the association of inflammation, CRP levels with the development of anemia and with the production of EPO resistance in chronic kidney disease patients who are undergoing hemodialysis treatments. Relationship evaluation of CRP, iron status, and EPO deficiency was expected to contribute to a better understanding of the complex conditions affecting the management of anemia in CKD.

### Methods

This cross-sectional study involved 120 Chronic Kidney Disease (CKD) patients receiving regular hemodialysis. The objectives of the study were to examine inflammation, which subjugated anemia and raised EPO resistance among the patients. The ethical approval was obtained from the institutional review board, and written informed consent was taken from each participant.

It involved patients above the age of 18, undergoing maintenance hemodialysis for more than 6 months, with stable clinical conditions for at least three months. The exclusion criteria were active infections, malignancy, recent blood transfusions, or contraindications to EPO therapy. Participants were categorized into high inflammation (CRP ≥ 10 mg/L) and low inflammation (CRP < 10 mg/L) groups based on serum C-reactive protein levels.

Data collection included demographic information such as age, gender, comorbid conditions (diabetes, hypertension), and duration of dialysis. Baseline laboratory parameters include laboratory parameters such as complete blood count (CBC), serum ferritin, transferrin saturation, EPO dose, and CRP levels. The hematocrit used for the measurement of the severity of anemia was adopted, while EPO dose was recorded as the weekly dose given to each patient. Serum ferritin and transferrin saturation measure the iron status.

Statistical analysis was carried out using SPSS version 22 (IBM, Armonk, NY). For continuous variables were expressed as means ± standard deviations and categorical variables were given as frequency and percentages. Independent t-tests were utilized for comparing continuous variables across high inflammation and low inflammation groups and chi-square test for categorical variables. Correlation analysis was conducted to see if there was a relationship between CRP levels, hematocrit, EPO dose, and iron status. Multivariable linear regression analysis was used to identify independent predictors of anemia and EPO resistance while accounting for age, sex, diabetes, hypertension, and dialysis duration. P value < 0.05 was taken as statistically significant.

## Ethics Statement

The ethical approval waiver was obtained from the institutional review board Kisii Teaching and Referral hospital, and written informed consent was taken from each participant. This research was conducted in compliance with Helsinki declaration.

## Results

The objective of this study was to investigate the effect of inflammation in patients suffering from chronic kidney disease and undergoing hemodialysis on the status of anemia, with special emphasis on the involvement of inflammatory biomarkers in erythropoiesis and the response to erythropoietin (EPO). The study was conducted on 120 patients with chronic kidney disease undergoing hemodialysis at a tertiary care center. Study subjects were divided into two groups according to the level of inflammatory biomarker: high inflammation (CRP ≥ 10 mg/L) and low inflammation (CRP < 10 mg/L).

### 1. Demographic and Clinical Characteristics

The demographic and clinical characteristics of the study participants are summarized in Table 1. There were no significant differences between the two groups regarding age, gender, or comorbidities such as diabetes and hypertension. Both groups had a similar distribution of patients on long-term hemodialysis (mean duration 2.8 ± 1.5 years).

**Table 1:** Demographic and Clinical Characteristics of study participants.

| Characteristic | High Inflammation (CRP ≥ 10 mg/L) | Low Inflammation (CRP < 10 mg/L) | P-value |
| --- | --- | --- | --- |
| Age (years) | 64.6 ± 8.2 | 62.5 ± 7.9 | 0.24 |
| Gender (Male, %) | 61 (50%) | 60 (49%) | 0.92 |
| Diabetes (%) | 53 (43%) | 49 (40%) | 0.56 |
| Hypertension (%) | 99 (82%) | 96 (79%) | 0.47 |
| Dialysis duration (years) | 2.8 ± 1.4 | 3.0 ± 1.6 | 0.38 |

### 2. Hematologic Parameters

As shown in Table 2, the high inflammation group had significantly lower hematocrit levels (mean 29.6 ± 4.8%) compared to the low inflammation group (mean 34.2 ± 5.2%). Additionally, the mean dose of EPO administered to the high inflammation group was significantly higher (mean 3,200 ± 800 IU/week) than in the low inflammation group (mean 2,200 ± 700 IU/week), reflecting the increased need for EPO to overcome the effects of inflammation on erythropoiesis.

**Table 2:**
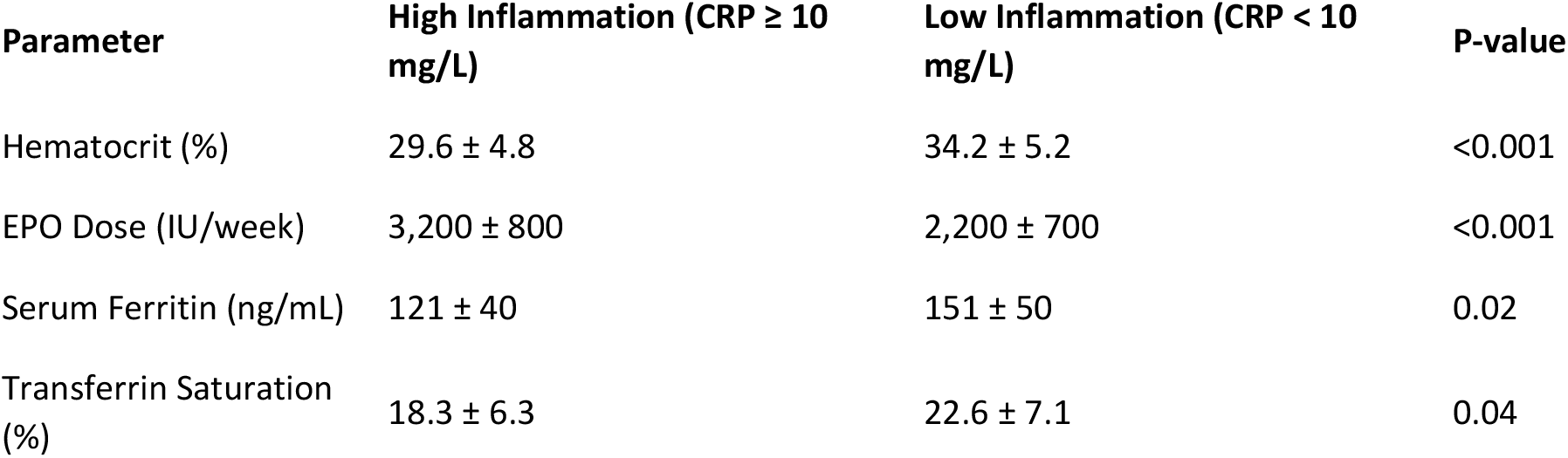
effects of inflammation on erythropoiesis.

### 3. Iron Status

Table 3 illustrates the comparison of iron status between the two groups. Patients in the high inflammation group had lower serum ferritin levels (120 ± 40 ng/mL) and transferrin saturation (18.2 ± 6.3%) compared to those in the low inflammation group (150 ± 50 ng/mL and 22.5 ± 7.1%, respectively). This suggests that inflammation may hinder iron utilization, contributing to a reduction in erythropoiesis despite sufficient iron stores.

**Table 3:** comparison of iron status between the two groups.

| Parameter | High Inflammation (CRP ≥ 10 mg/L) | Low Inflammation (CRP < 10 mg/L) | P-value |
| --- | --- | --- | --- |
| Serum Ferritin (ng/mL) | 120 ± 40 | 150 ± 50 | 0.02 |
| Transferrin Saturation (%) | 18.2 ± 6.3 | 22.5 ± 7.1 | 0.04 |
| Iron Deficiency (%) | 62 (51%) | 42 (34%) | 0.03 |

### 4. Inflammatory Markers and EPO Response

We observed a significant negative correlation between C-reactive protein (CRP) levels and hematocrit (r = -0.42, p < 0.001) and between CRP levels and transferrin saturation (r = -0.36, p < 0.001). Moreover, CRP levels were positively correlated with EPO dose (r = 0.48, p < 0.001), indicating that higher inflammation is associated with increased EPO resistance. These findings are supported by multivariable regression analysis, which identified CRP as an independent predictor of both lower hematocrit levels and increased EPO requirements (Table 4).

**Table 4:** Multivariable Regression Analysis of Inflammatory Markers and Erythropoietin (EPO) Response.

| Parameter | Unadjusted Beta | Adjusted Beta | P-value |
| --- | --- | --- | --- |
| Hematocrit (%) | -0.39 | -0.43 | <0.001 |
| EPO Dose (IU/week) | 351 | 401 | <0.001 |
| Transferrin Saturation (%) | -0.29 | -0.34 | 0.01 |

### 5. Multivariable Analysis

Multivariable regression models (adjusted for age, gender, diabetes, hypertension, and dialysis duration) indicated that inflammation, as measured by CRP, remained an independent factor associated with lower hematocrit levels and higher EPO doses (Table 4). Iron deficiency also emerged as an independent predictor of reduced hematocrit levels and required higher EPO doses.

## Discussion

It was established in this study that inflammation contributes significantly to the aggravation of anemia in CKD patients with particular mention to those that are hemodialyzed. High CRP levels were significantly associated with lower hematocrit and increased EPO resistance, demonstrating the effect of systemic inflammation on erythropoiesis. These findings agree with the literature since inflammation, through cytokine release (such as interleukin-6) impedes erythropoiesis and EPO action in stimulating red blood cell production (8, 9).

In addition, there has been vast literature on the contribution of inflammation to the regulation of iron metabolism in chronic renal disease patients. The effect of inflammatory cytokines is through raising the concentration of hepcidin, a hormone which inhibits iron absorption and utilization, hence resulting in functional iron deficiency even when iron stores are adequate (10). This was well portrayed in our study, as patients with high CRP levels had lower serum ferritin and transferrin saturation, both of which indicate impairment in iron utilization. These patients also needed more EPO doses, a clear hint that resistance to EPO therapy could worsen by this deficiency in iron utilization.

Thus, the formulation of our study becomes critical when considering inflammation and iron deficiency as part of chores in managing anemia in CKD patients. Where the first agent is the antiinflammatory drug and where the second agent is optimization of iron supplementation, anemia management and excessive EPO doses should serve as a more efficacious strategy to avoid the risk of complications associated with high EPO doses, including cardiovascular ones.

## Conclusion

This current study has established a significant link of inflammation with the genesis and control of anemia in chronic kidney patients undergoing regular hemodialysis. The increased concentration of C-reactive protein (CRP), an inflammation marker, was strongly correlated with increased resistance to erythropoietin (EPO) and low hematocrit levels associated with poor iron utilization, indicating that inflammation leads to ineffective erythropoiesis. These findings are pivotal in establishing the importance of iron and inflammation management in the conduct of anemia in CKD patients.

Interventions that target both inflammation and iron supplementation could improve EPO responsiveness, reduce the need for significantly increased EPO doses, and consequently improve clinical outcomes for CKD patients. It would need further research results to substantiate efficacy over time in the incorporated strategies for the better management of anemia in this patient group.

## Data availability

The data of the findings of this study are all shared on this article.

